# Laboratory and field evaluation of the STANDARD Q and Panbio™ SARS-CoV-2 antigen rapid test in Namibia using nasopharyngeal samples

**DOI:** 10.1101/2021.09.21.21263886

**Authors:** Iyaloo Konstantinus, Douglas Chiwara, Emmy-Else Ndevaetela, Victoria Ndarukwa-Phiri, Nathalia !Garus-oas, Ndahafa Frans, Pentikainen Ndumbu, Andreas Shiningavamwe, Gerhard van Rooyen, Ferlin Schiceya, Lindile Hlahla, Pendapala Namundjebo, Ifeoma Ndozi-Okia, Francis Chikuse, Sirak Hailu Bantiewalu, Kapena Tjombonde

## Abstract

**Background:** As new SARS-CoV-2 variants of concern emerge, there is a need to scale up testing to minimize transmission of the Coronavirus disease 2019 (COVID-19). Many countries especially those in the developing world continue to struggle with scaling up reverse transcriptase polymerase reaction (RT-PCR) to detect SARS-CoV-2 due to scarcity of resources. Alternatives such as antigen rapid diagnostics tests (Ag-RDTs) may provide a solution to enable countries to scale up testing.

**Methods:** In this study, we evaluated the Panbio™ and STANDARD Q Ag-RDTs in the laboratory using 80 COVID-19 RT-PCR confirmed and 80 negative nasopharyngeal swabs. The STANDARD-Q was further evaluated in the field on 112 symptomatic and 61 asymptomatic participants.

**Results:** For the laboratory evaluation, both tests had a sensitivity above 80% (Panbio™ = 86% vs STANDARD Q = 88%). The specificity of the Panbio™ was 100%, while that of the STANDARD Q was 99%. When evaluated in the field, the STANDARD Q maintained a high specificity of 99%, however the sensitivity was reduced to 56%.

**Conclusion:** Using Ag-RDTs in low resource settings will be helpful, however, negative results should be confirmed by RT-PCR where possible to rule out COVID-19 infection.

## Introduction

The pandemic caused by Severe Acute Respiratory Syndrome Coronavirus 2 (SARS-CoV-2) identified originally in China has now become a public health throughout the world, and we are now dealing with emerging variants of concern (VOCs) of which some are more infectious compared to the founder virus [1,2]. Reverse transcription polymerase chain reaction (RT-PCR) is the laboratory gold standard method to detect SARS-CoV-2 in people with coronavirus disease 2019 (COVID-19). However, this method has its challenges such as long turnaround time, high-cost and requires trained laboratory personnel.

Due to the challenges of using RT-PCR, antigen-rapid diagnostic tests (Ag-RDTs) are being considered in several countries for epidemiological surveillance and even diagnostic purposes in symptomatic individuals [3]. These tests are less expensive, produce results faster than molecular tests; yielding results in as little as 15 to 30 minutes, and they do not require specialized laboratory techniques [4,5]. The WHO recommended that Ag-RDTs that meet at least 80% sensitivity and 97% specificity can be utilized in settings where RT-PCR is limited [6]. The Namibian Medical Regulatory Council (NMRC) has proposed a framework for these tests to undergo in-country laboratory and field verification before being recommend for use.

In the present study, the Panbio™ and STANDARD Q were evaluated in detecting SARS-CoV-2 antigens in frozen nasopharyngeal samples The STANDARD Q test was further evaluated in the field using fresh nasopharyngeal samples.

## Materials and Methods

### Study design and population

The laboratory evaluation was a cross-sectional, retrospective verification of the performance of the STANDARD Q (SD Biosensor, Republic of Korea) and Panbio™ (Abbott Diagnostic GmbH). Nasopharyngeal samples from 80 SARS-CoV-2 positive and 80 SARS-CoV-2 negative RT-PCR confirmed participants were used for this evaluation. The field evaluation was a prospective cross-sectional, prospective verification of the STANDARD Q Ag-RDT. Participants 18 years and older seeking SARS-CoV-2 testing were recruited from the Robert Mugabe Clinic and the Katutura Health Center. Participants were either symptomatic with flu-like symptoms, or asymptomatic cases who reported to have been high risk contacts of confirmed cases. This evaluation was approved by the research ethics committee of the Ministry of Health and Social Services (Ref:17/3/3/EEN). Samples were not linked to any personal identifier and results could not be traced back to individual patients.

### RT-PCR and Ag-RDT testing

Nasopharyngeal swabs collected in viral transport medium (VTM) were processed within 24 hours of collection. The viral load was expressed as cycle threshold (CT) value, and a cut of <40 was used. For the laboratory evaluation, swabs were stored at -80°C and retrieved for antigen testing within 48 hours by a laboratory personnel according to the manufacturer instructions. For the STANDARD Q, results were read strictly within 15-30 minutes, while for the Panbio™ results were read in 15 minutes and not beyond 20 minutes as per manufacturer’s instructions. For the field verification using the STANDARD Q, two nasopharyngeal swabs were collected; one performed by the clinicians on site on one swab, while the other was transported to the laboratory immediately for testing with RT-PCR.

### Statistical analysis

Sensitivity was calculated as the number of positive samples identified as positive for each Ag-RDT divided by the number of positive samples identified by RT-PCR reference assay. Specificity was calculated as the number of negative samples identified as negative for each Ag-RDT divided by the number of negative samples identified by RT-PCR reference assay. The Positive Predictive Values (PPV) and Negative Predictive Values (NPV) were also calculated. Mann-Whitney U test was computed to compare the differences between two groups using Prism V9 (GraphPad Software).

## Results

### Laboratory performance of the Panbio™ and STANDARD Q

To determine the specificity and sensitivity of both Ag-RDT, frozen VTM from nasopharyngeal swabs were used. The sensitivity for the STANDARD Q was 88% (95% CI: 79% to 93%) and the specificity was 99% (95% CI: 93% to 99%), while that of Panbio™ was 86% (95% CI: 76% to 91%) and 100% (95% CI: 95% to 100%) respectively (Table 1). The PPV for the STANDARD Q and Panbio™ was 99% and 100% respectively, while the NPV was 89% and 87% respectively.

**Table 1.** Determination of the Ag-RDTs sensitivity and specificity

| Ag-RDTs |  | RT-PCR |  | Total | Sensitivity | Specificity |
| --- | --- | --- | --- | --- | --- | --- |
| STANDARD Q |  | Positive | Negative |  | 88% | 99% |
|  | Positive | 70 | 1 | 71 |  |  |
|  | Negative | 10 | 79 | 89 |  |  |
|  | Total | 80 | 80 | 160 |  |  |
|  |  |  |  |  | 86% | 100% |
| Panbio™ | Positive | 68 | 0 | 68 |  |  |
|  | Negative | 12 | 79 | 91 |  |  |
|  | Total | 80 | 79 | 159 |  |  |

We then compared samples which were SARS-CoV-2 positive using RT-PCR grouped according to their Ag-RDT results (**Figure 1**). STANDARD Q had 70 true positives and 10 false negatives; and Panbio™ had 68 true positives and 12 false negatives. Interestingly for both Ag-RDTs, samples which had a negative result were those with a high CT value. For STANDARD Q, the median CT value for the negative samples was 33 compared to a median CT value of 24 in the positive samples. The median CT value for the negative samples using the Panbio™ was 32 compared to that of 24 in the positive samples.

**Figure 1.**
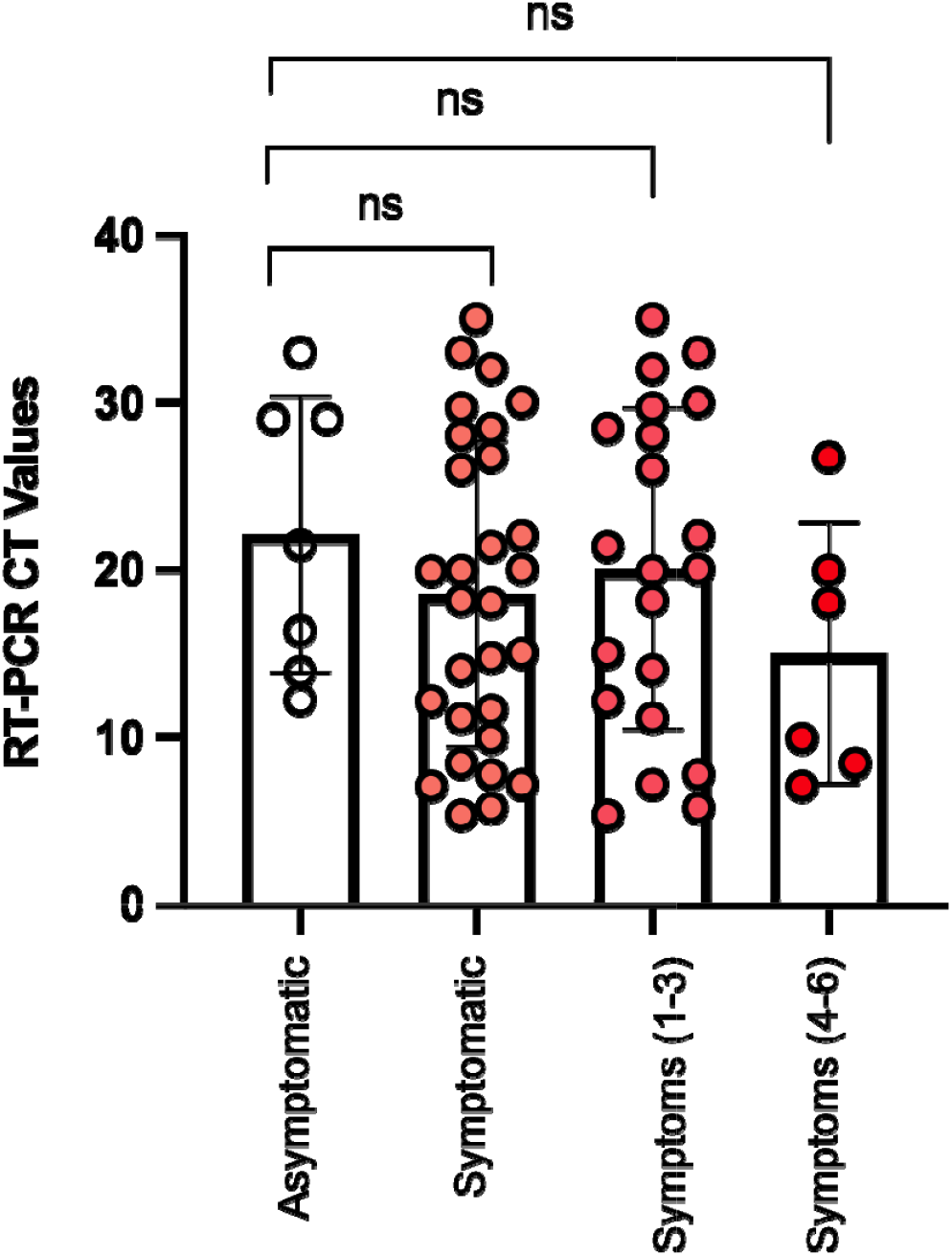
CT values in participants who tested positive for SARS-CoV-2 using RT-PCR, grouped according to each Ag-RDT positive and negative results (^****^p<0.0001).

### Characteristics of participants included in the field evaluation

Nasopharyngeal samples from 173 participants were included in the field evaluation. The demographic and clinical characteristics are shown in Table 2. The median age was 33 years, with an almost equal distribution of females (51%) and males (49%). Of these participants, 65% (n=112) reported having symptoms onset within 5-7 days, with having a cough being the most prevalent symptom (37%) followed by headache (28%); while vomiting was the least common symptom in this cohort. A total of 36 samples tested positive by RT-PCR representing 21% positivity, of which 29 samples were from symptomatic and 7 were from asymptomatic participants.

**Table 2.** Demographics and clinical characteristics of participants

|  | All<br>n=173 | RT-PCR +<br>n=36 | RT-PCR –<br>n=137 |
| --- | --- | --- | --- |
| Age, Median [IQR] | 33 [23, 44] | 39 [25, 44] | 32 [23, 44] |
| Sex |  |  |  |
| Female | 88 (51) | 19 (53) | 69 (50) |
| Male | 85 (49) | 17 (47) | 68 (50) |
| Symptomatic |  |  |  |
| No | 61 (35) | 7 (19) | 54 (39) |
| Yes | 112 (65) | 29 (81) | 83 (61) |
| Symptoms |  |  |  |
| Cough | 64 (37) | 18 (50) | 46 (34) |
| Fever | 12 (7) | 2 (6) | 10 (7) |
| Sore throat | 35 (20) | 8 (22) | 27 (20) |
| Diarrhoea | 9 (5) | 1 (3) | 8 (6) |
| Loss of smell | 16 (9) | 5 (14) | 11 (8) |
| Chills | 15 (9) | 5 (14) | 10 (7) |
| Shortness of breath | 5 (3) | 0 (0) | 5 (4) |
| Myalgia | 12 (7) | 5 | 7 (5) |
| Vomiting | 1 (1) | 1 | 0 (0) |
| Loss of taste | 20 (12) | 6 | 14 (10) |
| Runny nose | 27 (16) | 10 | 17 (12) |
| Headache | 49 (28) | 14 | 35 (26) |
| Blocked nose | 8 (5) | 0 | 8 (6) |
| Chest pain | 11 (6) | 0 | 11 (8) |
| CT value, Mean [Range] | 19 [5, 30] |  |  |
| CT, Cycle threshold; IQR, Interquartile range; RT-PCR, Reverse transcriptase polymerase reaction |  |  |  |

**Table 3.** Field sensitivity and specificity of the STANDARD-Q Ag-RDT

| Ag-RDT |  | RT-PCR |  | Total | Sensitivity | Specificity |
| --- | --- | --- | --- | --- | --- | --- |
| STANDARD Q<br>COVID-19 Ag |  | Positive | Negative |  | 56% | 99% |
|  | Positive | 20 | 1 | 21 |  |  |
|  | Negative | 16 | 136 | 152 |  |  |
|  | Total | 36 | 137 | 173 |  |  |

### STANDARD Q Ag-RDT had a decreased sensitivity in the field

For the field evaluation, only the STANDARD Q Ag-RDT was evaluated due to the unavailability of more Panbio™ kits. The STANDARD Q test had a decreased sensitivity of 56% (95% CI: 40% vs 70%) compared to the laboratory sensitivity of 87%. The specificity in the field was high at 99% (95% CI: 96% to 99%) similar to that observed in the laboratory. The PPV was 95%, while the NPV was 90%.

We also compared the CT values in participants who were SARS-CoV-2 PCR positive grouped according to their STANDARD Q test result (**Figure 2A**). Participants with concordant positive results (RT-PCR+ STANDARD Q+) had a median CT value of 13 (IQR: 8, 19) while those with discordant results (RT-PCR+ STANDARD Q-) had a median CT value of 28 (IQR: 20, 30). We further grouped these participants according to symptoms; asymptomatic, all symptomatic, those who had 1 to 3 symptoms, and those who had 4-6 symptoms. Although the was a trend towards a high median CT value in asymptomatic compared to symptomatic participants, this was not significant (**Figure 2B**).

**Figure 2.**
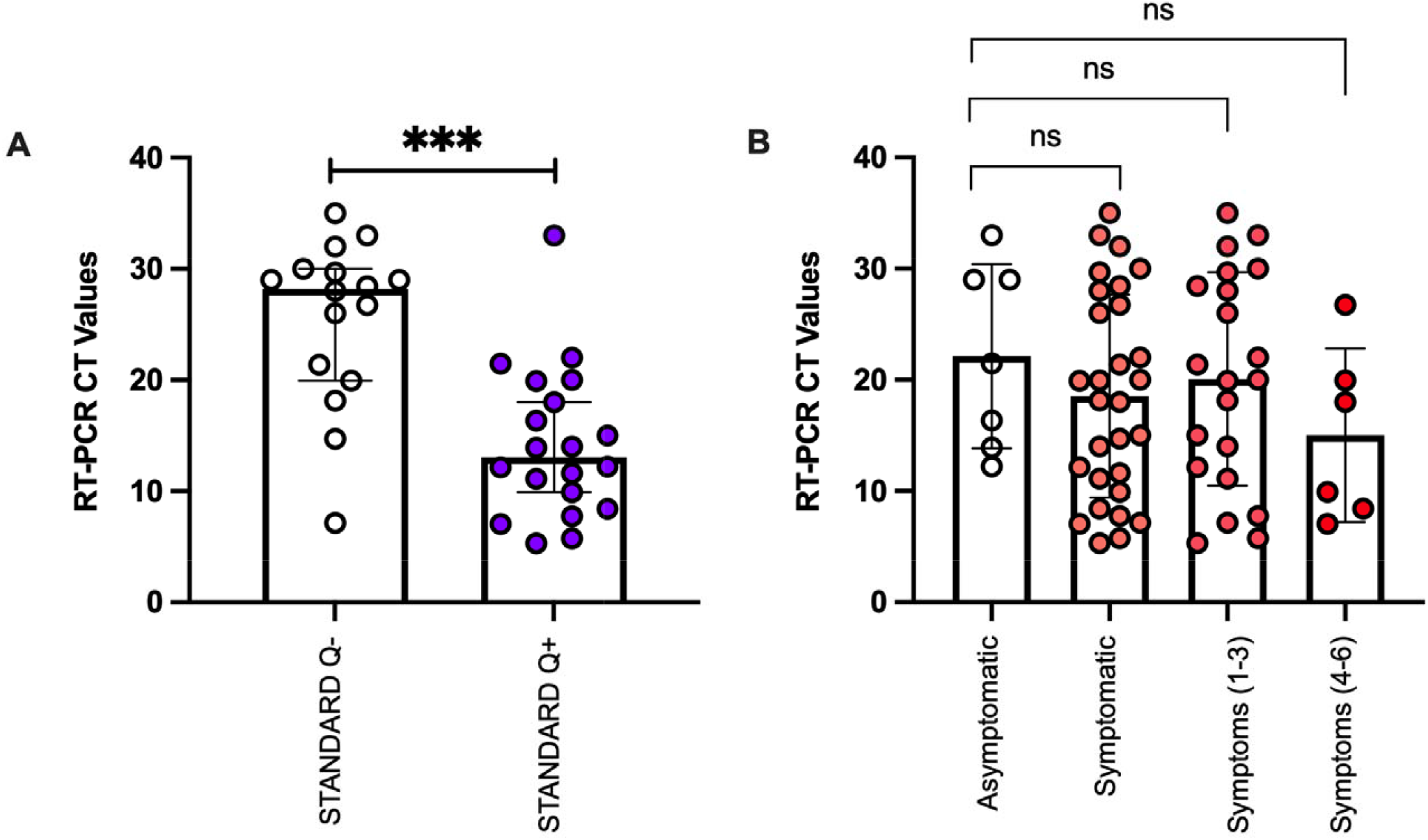
CT values in participants who tested positive for SARS CoV-2 by RT-PCR. (A) Participants grouped according to the STANDARD Q results (***p=0.0001). (B) Participants were grouped as asymptomatic, all symptomatic, having 1 to 3 symptoms and having 4 to 6 symptoms. P-value was not significant (ns).

## Discussion

Molecular testing is inherently difficult to scale up as it requires laboratories with specialized equipment and reagents which are costly, and trained laboratory personnel [4,7]. By the end of 2020, African countries have seen an increase in COVID-19 cases in the second wave of the pandemic compared to the first wave, as new variants of concern such as the Beta and Delta reported to be more infectious spread across the continent [8,9]. Namibia has not been spared from this, being one of the worst affected African countries during the third wave. Timely and accurate COVID-19 testing is a critical component of surveillance, contact tracing, infection prevention and control and clinical management of COVID-19 cases. Hence, there is a need to scale up testing and Ag-RDTs might be useful for this in resource limited countries including Namibia.

In this study, we evaluated two Ag-RDTs, Panbio™ and STANDARD Q. The laboratory evaluation showed a high specificity for both tests, with the Panbio™ at 100% compared to the STANDARD Q at 98%. In contrast, the sensitivity of the STANDARD Q was slightly higher at 88% compared to that of the Panbio™ at 86%. For the field evaluation, only the STANDARD Q Ag-RDT was evaluated due to its availability at the time of conducting the study. Although the STANDARD Q maintained a high specificity of 99% in the field, the test had a reduced sensitivity of 56%. The decreased field sensitivity is expected because the laboratory evaluation used selected SARS-CoV-2 positive samples compared to testing participants in the field with unknown SARS-CoV-2 status and based on their self-reported symptoms. Hence, the performance of these tests also depends on the settings they are being used and the prevalence of the disease at the time of the study.

Several studies have reported Ag-RDTs to be more sensitive in samples with low CT values as a result of high viral load [10–14]. This was also observed in our evaluation for both Ag-RDTs. In the laboratory evaluation, the false negative samples were similar for both STANDARD Q and Panbio™ with the later having an additional two samples, and these samples had a CT value above 25. One study reported sensitivity of the Panbio™ and STANDARD Q to be a 100% for samples with a CT value below 20, decreasing at 41% and 52% respectively for those samples with a CT value between 25-30 [14]. Other studies also reported the sensitivity of these two Ag-RDTs to increase above 80% when the CT value is <25 [7,12,13,15–19].

One of the limitations for this study included the credibility of the symptom onset of the participants included in the field of evaluation. We could not determine with certainty whether the samples collected were from patients whose symptoms onset was within 5-7 days. This is important because these tests have been reported to be more reliable in detecting SARS-CoV-2 infection within the first 7 days after the onset of symptoms. Therefore, they can miss individuals who are in the very early stage of infection (presymptomatic stage) and those who are in the late stage with a decreased viral replication.

In conclusion, these results add to the body of evidence that Ag-RDTs are useful and may be utilized to scale up testing to reduce viral transmission in settings where RT-PCR is a challenge.

## Data Availability

Data is available on request

## Acknowledgements

We would like to thank the NIP COVID-19 Laboratory team, Dr Suzanne Beard from CDC, the Laboratory pillar at the Ministry of Health and Social Services (MoHSS) specifically Mrs Mary Mataranyika, and the staff at the Robert Mugabe and Katutura Health Centre.

## Authors contribution

Drafting of the manuscript: IK

Methodology design: DC, IK, AS, EN, VNP, FC, GvR, PN, INO, SHB

Sample collection: NG, NF, FS, LH, IK, DC, EN, VNP

Investigation: IK, EN, VNP

Wet lab experiments: NG, NF

Data capturing: IK

Data analysis: IK, DC, VNP

Reviewing and editing: IK, DC, KT, VNP, PN, NG

## References

1. Zhu N, Zhang D, Wang W, Li X, Yang B, Song J, et al. A novel coronavirus from patients with pneumonia in China, 2019. N Engl J Med. 2020;382: 727–733. doi:10.1056/NEJMoa2001017

2. Karim SSA, de Oliveira T. Correspondence New SARS-CoV-2 Variants — Clinical, Public Health, and Vaccine Implications. N Engl J Med. 2021; NEJMc2100362.

3. Abdelrazik AM, Elshafie SM, Abdelaziz HM. Potential Use of Antigen-Based Rapid Test for SARS-CoV-2 in Respiratory Specimens in Low-Resource Settings in Egypt for Symptomatic Patients and High-Risk Contacts. Lab Med. 2021;52: e46–e49. doi:10.1093/labmed/lmaa104

4. Peeling RW, Olliaro PL, Boeras DI, Fongwen N. Scaling up COVID-19 rapid antigen tests: promises and challenges. Lancet Infect Dis. 2021;3099: 21–26. doi:10.1016/S1473-3099(21)00048-7

5. Vandenberg O, Martiny D, Rochas O, van Belkum A, Kozlakidis Z. Considerations for diagnostic COVID-19 tests. Nat Rev Microbiol. 2021;19: 171–183. doi:10.1038/s41579-020-00461-z

6. WHO. Antigen-detection in the diagnosis of SARS-CoV-2 infection using rapid immunoassays Interim guidance, 11 September 2020. World Heal Organ. 2020; 1–9. Available: https://www.who.int/publications/i/item/antigen-detection-in-the-diagnosis-of-sars-cov-2infection-using-rapid-immunoassays%0Ahttps://apps.who.int/iris/handle/10665/334253

7. Cerutti F, Burdino E, Milia MG, Allice T, Gregori G, Bruzzone B, et al. Urgent need of rapid tests for SARS CoV-2 antigen detection: Evaluation of the SD-Biosensor antigen test for SARS-CoV-2. J Clin Virol. 2020;132: 104654. doi:10.1016/j.jcv.2020.104654

8. Jassat W, Mudara C, Ozougwu L, Tempia S, Blumberg L, Davies M-A, et al. Difference in mortality among individuals admitted to hospital with COVID-19 during the first and second waves in South Africa: a cohort study. Lancet Glob Heal. 2021; 1–10. doi:10.1016/S2214-109X(21)00289-8

9. Salyer SJ, Maeda J, Sembuche S, Kebede Y, Tshangela A, Moussif M, et al. The first and second waves of the COVID-19 pandemic in Africa: a cross-sectional study. Lancet. 2021;397: 1265–1275. doi:10.1016/S0140-6736(21)00632-2

10. Favresse J, Gillot C, Oliveira M, Cadrobbi J, Elsen M, Eucher C, et al. Head-to-Head Comparison of Rapid and Automated Antigen Detection Tests for the Diagnosis of SARS-CoV-2 Infection. J Clin Med. 2021;10: 265. doi:10.3390/jcm10020265

11. Gremmels H, Winkel BMF, Schuurman R, Rosingh A, Rigter NAM, Rodriguez O, et al. Real-life validation of the Panbio™ COVID-19 antigen rapid test (Abbott) in community-dwelling subjects with symptoms of potential SARS-CoV-2 infection. EClinicalMedicine. 2021;31: 100677. doi:10.1016/j.eclinm.2020.100677

12. Fenollar F, Bouam A, Ballouche M, Fuster L, Prudent E, Colson P, et al. Evaluation of the panbio COVID-19 rapid antigen detection test device for the screening of patients with COVID-19. J Clin Microbiol. 2020;59: 19–21. doi:10.1128/JCM.02589-20

13. Albert E, Torres I, Bueno F, Huntley D, Molla E, Fernández-Fuentes MÁ, et al. Field evaluation of a rapid antigen test (Panbio™ COVID-19 Ag Rapid Test Device) for COVID-19 diagnosis in primary healthcare centres. Clin Microbiol Infect. 2020;27: 472.e7–472.e10. doi:10.1016/j.cmi.2020.11.004

14. Pérez-García F, Romanyk J, Moya Gutiérrez H, Labrador Ballestero A, Pérez Ranz I, González Arroyo J, et al. Comparative evaluation of Panbio and SD Biosensor antigen rapid diagnostic tests for COVID-19 diagnosis. J Med Virol. 2021;93: 5650–5654. doi:10.1002/jmv.27089

15. Lanser L, Bellmann-Weiler R, Öttl KW, Huber L, Griesmacher A, Theurl I, et al. Evaluating the clinical utility and sensitivity of SARS-CoV-2 antigen testing in relation to RT-PCR Ct values. Infection. 2021;49: 555–557. doi:10.1007/s15010-020-01542-0

16. Mak GC, Lau SS, Wong KK, Chow NL, Lau CS, Lam ET, et al. Analytical sensitivity and clinical sensitivity of the three rapid antigen detection kits for detection of SARS-CoV-2 virus. J Clin Virol. 2020;133: 104684. doi:10.1016/j.jcv.2020.104684

17. Iglòi Z, Velzing J, Van Beek J, Van de Vijver D, Aron G, Ensing R, et al. Clinical evaluation of roche sd biosensor rapid antigen test for sars-cov-2 in municipal health service testing site, the netherlands. Emerg Infect Dis. 2021;27: 1323–1329. doi:10.3201/eid2705.204688

18. Boum Y, Fai KN, Nicolay B, Mboringong AB, Bebell LM, Ndifon M, et al. Performance and operational feasibility of antigen and antibody rapid diagnostic tests for COVID-19 in symptomatic and asymptomatic patients in Cameroon: a clinical, prospective, diagnostic accuracy study. Lancet Infect Dis. 2021. doi:10.1016/S1473-3099(21)00132-8

19. Jääskeläinen AE, Ahava MJ, Jokela P, Szirovicza L, Pohjala S, Vapalahti O, et al. Evaluation of three rapid lateral flow antigen detection tests for the diagnosis of SARS-CoV-2 infection. J Clin Virol. 2021;137: 104785. doi:https://doi.org/10.1016/j.jcv.2021.104785

